# A comparison of diffusion MRI presurgical tractography techniques with intraoperative mapping-based validation

**DOI:** 10.1101/2023.06.13.23290806

**Authors:** A.M. Radwan, L. Emsell, K. Vansteelandt, E. Cleeren, R. Peeters, S. De Vleeschouwer, T. Theys, P. Dupont, S. Sunaert

## Abstract

**Objectives:** Accurate presurgical brain mapping enables preoperative risk assessment and intraoperative guidance. This work investigated whether constrained spherical deconvolution (CSD) methods were more accurate than diffusion tensor imaging (DTI)-based methods for presurgical white matter mapping using intraoperative direct electrical stimulation (DES) as the ground truth.

**Material and methods:** Five different tractography methods were compared (3 DTI-based and 2 CSD-based) in 22 preoperative neurosurgical patients. The corticospinal tract (CST, N=20) and arcuate fasciculus (AF, N=7) bundles were reconstructed, then minimum distances between tractograms and DES coordinates were compared between tractography methods. Receiver-operating characteristic (ROC) curves were used for both bundles. For the CST, binary agreement, linear modeling, and posthoc testing were used to compare tractography methods while correcting for relative lesion and bundle volumes.

**Results:** Distance measures between 154 positive (functional response, pDES) and negative (no response, nDES) coordinates, and 134 tractograms resulted in 860 data points. Higher agreement was found between pDES coordinates and CSD-based compared to DTI-based tractograms. ROC curves showed overall higher sensitivity at shorter distance cutoffs for CSD (8.5 mm) compared to DTI (14.5 mm). CSD-based CST tractograms showed significantly higher agreement with pDES, which was confirmed by linear modeling and posthoc tests (PFWE < 0.05).

**Conclusion:** CSD-based CST tractograms were more accurate than DTI-based ones when validated using DES-based assessment of motor and sensory function. This demonstrates the potential benefits of structural mapping using CSD in clinical practice.

**Clinical relevance statement:** CSD-based tractograms of the CST are more sensitive than DTI-based tractograms when validated against sensory-motor DES mapping. This also demonstrated the feasibility of fully-automated CSD-based tractography for presurgical planning of the CST.

**Graphical abstract:** Presurgical white matter mapping using probabilistic CSD tractography is more accurate and sensitive than manual DTI FACT or automated probabilistic DTI tractography. This study included 22 patients with DES data, which was used as the ground truth. Distance in mm between tractograms and DES data resulted in 860 datapoints, 685 of which belonged to the CST and were used for linear modeling, DTI = diffusion tensor imaging, CSD = constrained spherical deconvolution, TCK = tractogram/tractography, FWE = family-wise error rate, AUC = area under the curve

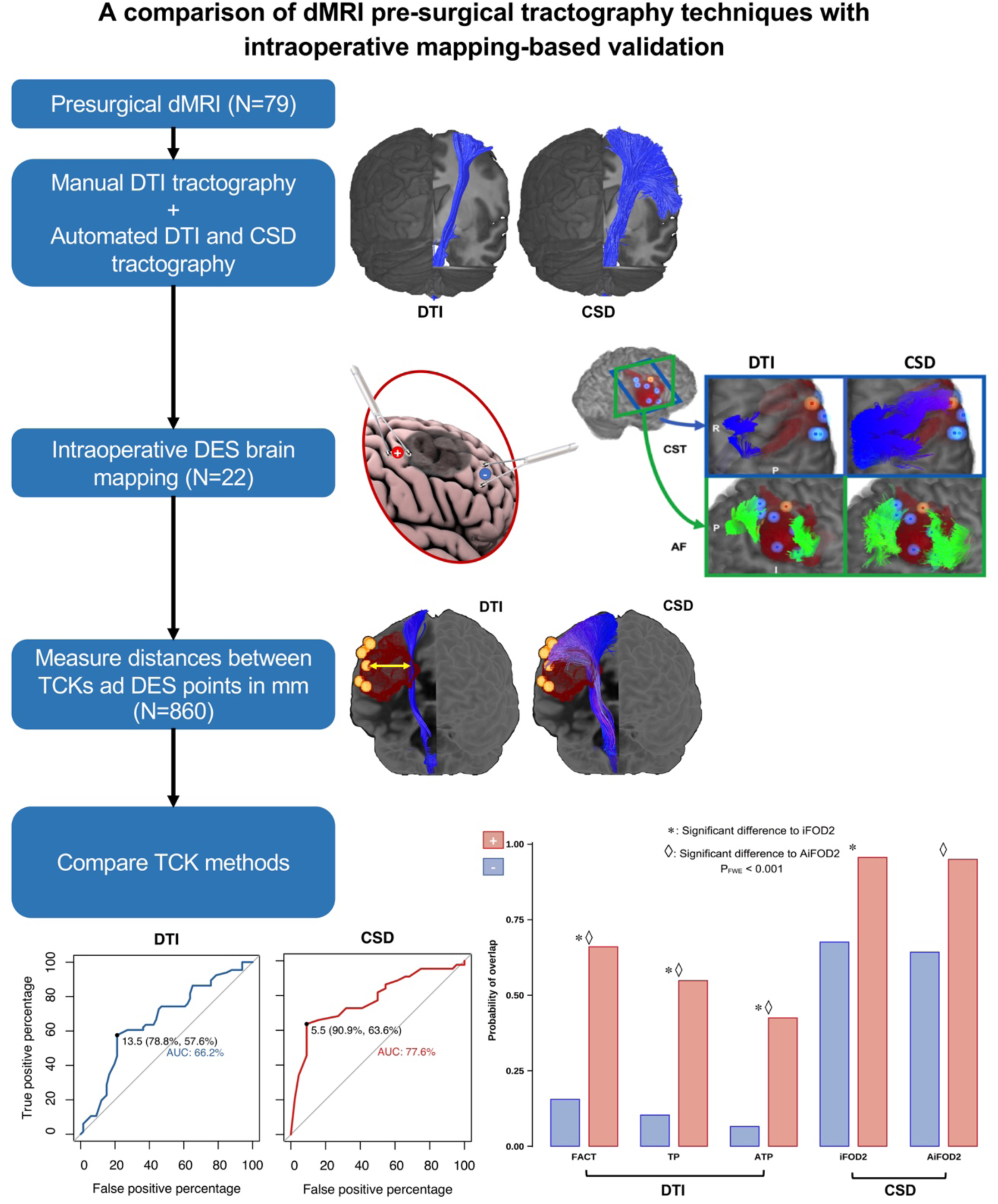

## Introduction

Modern neurosurgery strives to optimize functional preservation and therapeutic outcomes. The gold-standard for function-preserving brain surgery[1] is intraoperative direct electrical stimulation (DES) during awake-surgery with image-guided neuronavigation. Magnetic resonance imaging (MRI)-based non-invasive brain mapping, normally complements and guides DES[2–5]. However, it may be the sole mapping method available in patients where the gold-standard is not feasible due to its complexity and potential complications[6–8].

Here we focused on presurgical mapping with diffusion MRI (dMRI)-based fiber tractography (FT). FT is the rendering of white matter fasciculi in 3D using dMRI signal contrast and computational modeling[9]. Due to its clinical accessibility and validation[10–14], the most commonly used FT approach in neurosurgical settings is diffusion tensor imaging (DTI)-based fiber assignment by continuous tracking (FACT)[15].

DTI-FACT suffers from multiple limitations making it suboptimal in neurosurgical practice. DTI assumes a single fiber direction per voxel, which compromises the accuracy of modeled streamline trajectories in voxels containing complex fiber architecture, leading to underestimation of the true extent of fiber bundles[16, 17]. DTI-FACT is also confounded by lesion-effects, as perilesional edema can compromise sensitivity to fiber orientation, resulting in false-negatives, i.e., missing streamlines/tracts. Additionally, FT suffers from user-bias and limited reproducibility when manual region of interest (ROIs) definition is used, which is the most common approach in clinical practice. These short-comings limit the accuracy of presurgical fiber tracking and motivate exploring alternative methods using automation, and higher-order model based tractography[16].

High angular-resolution imaging (HARDI) methods like constrained spherical deconvolution (CSD)[18] can address the limitations of DTI-FACT[19–25]. However, HARDI typically requires a more complex data acquisition compared to DTI[26–29]. Some techniques, such as CSD have been shown to improve tractography based on single-shell low angular-resolution clinical data[30, 31]. Probabilistic-FT tends to have a higher sensitivity compared to deterministic-FT, and thus may be preferable in a surgical setting where false-negatives are more critical. Anatomically-constrained tractography (ACT)[32] may also improve accuracy of clinical tractography by constraining streamline origin/termination to the gray-white matter interface.

In this study we compared distance measures between DES coordinates acquired intraoperatively through pathology-tailored craniotomies and tractograms generated with deterministic DTI-FACT, probabilistic-DTI, and probabilistic-CSD, with and without ACT, to determine their respective suitability for presurgical mapping. We focused on the corticospinal tract (CST) and arcuate fasciculus (AF) given their relevance in clinical practice as primary pathways for sensory-motor and language functions.

## Methodology

### Research questions

We investigated the following research questions (RQ) in order to evaluate the performance of CSD-and DTI-based tractography with DES as the ground truth:

**RQ1:** How do distance measures between tractograms and DES coordinates compare between different tractography methods?

**RQ2:** How do these methods compare in binary agreement/disagreement with DES at different distance cutoffs?

**RQ3:** Are differences between tractography methods in the CST dependent on bundle volume and/or lesion volume?

### Participants

We recruited 79 surgery-naïve patients referred for presurgical fMRI and DTI between 01/2019 and 01/2021, 22 patients also underwent intraoperative DES mapping (14 males, age=8 – 73 years, median=39.5, IQR=28, Neoplasms=18 and focal cortical dysplasia=4). Each patient was informed about the study and signed a written informed consent before participation, in accordance with the declaration of Helsinki. Local ethics committee approval was obtained (UZ/KU Leuven, Leuven, Belgium, S61759). Participating patients were excluded if they had undergone previous resective brain surgery, had brain implants, ventriculoperitoneal shunts, or had absolute contraindications to MRI. Detailed demographics and pathological information can be found in **S.table 1**.

**Table 1:** MRI acquisition parameters.

| Table 1: MRI acquisition parameters |  |  |  |
| --- | --- | --- | --- |
|  | dMRI - part1 | dMRI - part2 | dMRI - part3 |
| Pulse sequence | 2D spin-echo EPI |  |  |
| Acquisition plane | Axial |  |  |
| TR/TE ms | 4500/85 |  |  |
| FA ° | 85 |  |  |
| Voxel size in mm<sup>3</sup> | 1.96*1.96*2.00 |  |  |
| Acquisition matrix | 112*112*69 |  |  |
| In-plane acceleration | SENSE 1.6 |  |  |
| Multiband | 3 |  |  |
| Pixel BW | 2997 |  |  |
| PE direction | AP |  |  |
| Fat shift direction | A |  | P |
| Bvalue (s/mm<sup>2</sup>) | 1200 | 2500 | 0 |
| Number of diffusion directions | 127 | 124 | 0 |
| Number of non-diffusion volumes | 1 | 4 - 9 | 4 - 7 |
| dMRI = diffusion magnetic resonance imaging, EPI = echo planar imaging, TR = repetition time, TE = echo time, ms = milliseconds, FA = flip angle, SENSE = SENSitivity encoding in plane parallel imaging acceleration, BW = bandwidth, PE = phase encoding, AP = anteroposterior, A = anterior, P = posterior |  |  |  |

### MRI acquisition

Two 3-tesla MRI scanners were used for multimodal presurgical MRI scanning (Ingenia - Elition, and Achieva DStream, Philips Medical Systems, Best, The Netherlands), both with 32-channel receive head coils. The acquisition parameters for 3D T1-weighted images, T2-and T2 fluid attenuation inversion recovery (FLAIR) images were previously described [33]. Multi-shell dMRI data and reversed phase B0 images were acquired whenever feasible and tolerated by the patient, see **Table 1**. Single-shell data with or without reversed phase B0 images were used if multi-shell data couldn’t be acquired, see **S.table 2**.

**Table 2:** Summarized binary agreement/disagreement confusion matrix results for all Tractogram-DES pairs.

|  |  |  |  |  |  |  |
| --- | --- | --- | --- | --- | --- | --- |
| FT methods | Thresholds (mm) | Accuracy | Sensitivity | Specificity | Pos Pred Value | Neg Pred Value |
| FACT | 5.5 | 75% | 33% | 92% | 63% | 77% |
|  | 12.5 | 71% | 61% | 75% | 50% | 82% |
|  | 13.5 | 71% | 65% | 74% | 51% | 83% |
| TP | 5.5 | 68% | 39% | 80% | 45% | 76% |
|  | 12.5 | 70% | 67% | 72% | 49% | 84% |
|  | 13.5 | 70% | 67% | 71% | 49% | 84% |
| ATP | 5.5 | 69% | 25% | 87% | 45% | 74% |
|  | 12.5 | 72% | 57% | 78% | 52% | 81% |
|  | 13.5 | 72% | 59% | 77% | 52% | 82% |
| iFOD2 | 5.5 | 74% | 65% | 77% | 54% | 84% |
|  | 12.5 | 57% | 88% | 45% | 40% | 90% |
|  | 13.5 | 57% | 90% | 43% | 40% | 91% |
| AiFOD2 | 5.5 | 75% | 71% | 77% | 56% | 86% |
|  | 12.5 | 60% | 92% | 47% | 42% | 94% |
|  | 13.5 | 58% | 92% | 44% | 41% | 93% |
| FT = fiber tractography, Pos Pred = positive predictive, Neg Pred = negative predictive |  |  |  |  |  |  |

### Intraoperative brain mapping

Twenty patients underwent awake neurosurgery and DES, and 2 patients underwent DES with motor and somatosensory evoked potentials (MEP/SSEP). Intraoperative frameless neuronavigation (Curve, BrainLab, Munich, Germany) was employed in all cases. DES used the OSIRIS neurostimulator (Inomed Medizintechnik GmbH, Germany), and a bipolar fork stimulator with 5mm inter-electrode spacing for cortical mapping, and a monopolar suction-stimulator for subcortical mapping.

### Data analysis

**Figure 1** shows a schematic representation of the data preprocessing and analysis workflow. All acquired images were converted to the brain imaging data structure format (BIDS) [34] format using the KU Leuven Neuroimaging suite (KUL_NIS) [35] (https://github.com/treanus/KUL_NIS) and dcm2bids [36].

**Figure 1:**
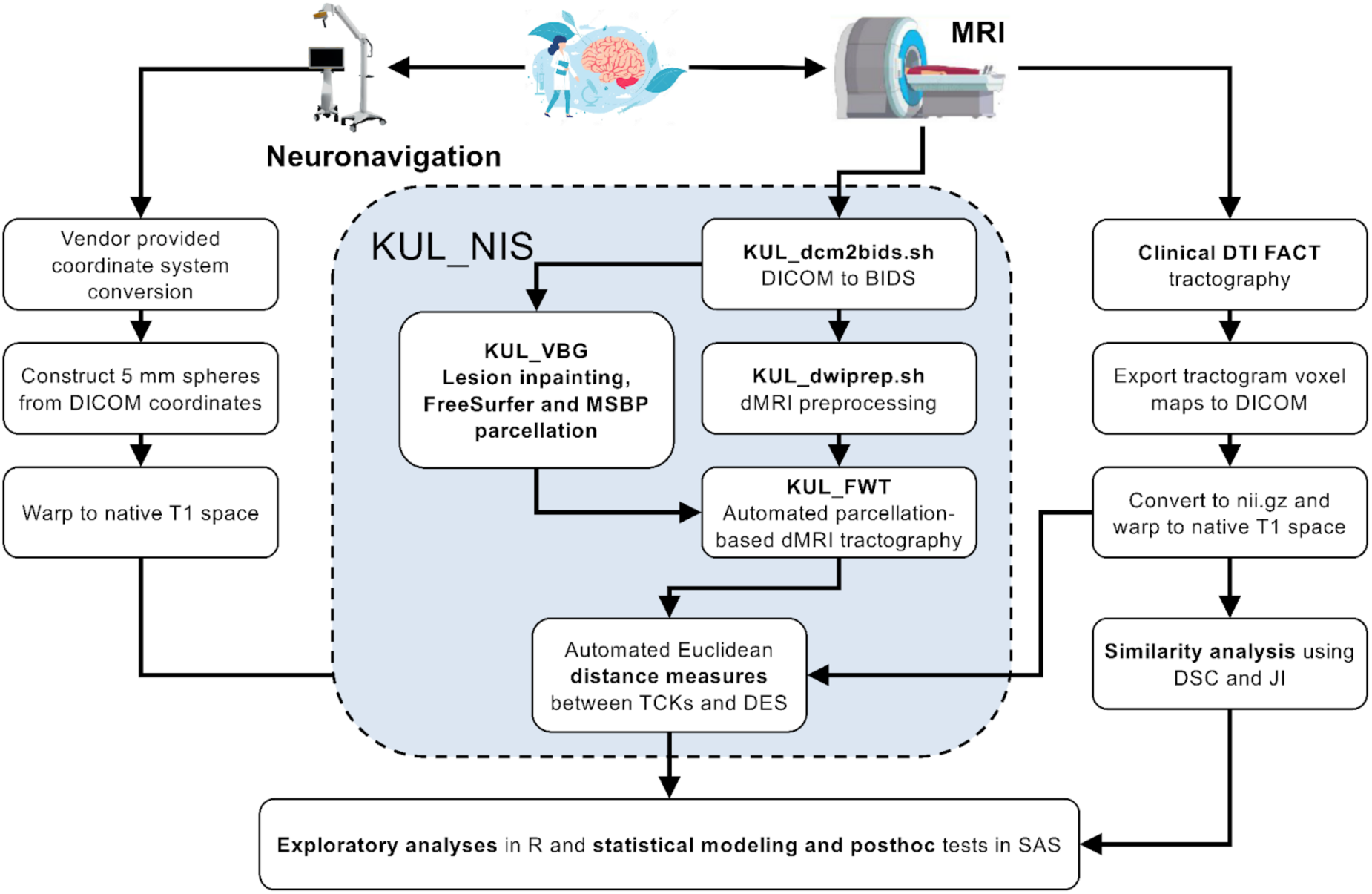
Schematic representation of the data preprocessing and analysis workflow used to compare different tractography results to intraoperative mapping outcome. MRI = magnetic resonance imaging, KUL_NIS = KU Leuven neuroimaging suite, BIDS = brain imaging data structure, KUL_VBG = KU Leuven virtual brain grafting, KUL_FWT = KU Leuven fun with tracts, DTI = diffusion tensor imaging, FACT = fiber assignment by continuous tracking, DSC = dice similarity coefficient, JI = Jaccard index

### Structural images

Semi-automated classification in ITK-snap v3.8.0[37] was used for lesion segmentation using T1, T2, T2-FLAIR and contrast-enhanced T1-weighted images. KU Leuven Virtual brain grafting v0.52 (VBG)[33] (https://github.com/KUL-Radneuron/KUL_VBG) was used for T1 lesion-filling and T1 parcellation[38–40]. FreeSurfer[39] generated 1mm isotropic T1-weighted images were used as base images for calculating distances. Lesion volume, total intracranial volume (TIV), and lesion mask to TIV ratios were also calculated per patient.

### Diffusion-weighted images

Routine clinical dMRI analysis involved in-line rigid interframe registration for motion correction on a clinical workstation (Philips Medical Systems, Best, The Netherlands). Advanced dMRI preprocessing used the KUL_dwiprep.sh script[35], which relies on FSL v6.0[41], ANTs v2.3.0[42, 43] and MRtrix3 v3.0.3[44] for denoising, correction of Gibbs ringing, subject motion, Eddy current artifacts, echo-planar imaging (EPI) distortion, and imaging bias. This was followed by DTI and CSD model fitting in MRtrix3[44–46]. FSL’s topup[47] was used for EPI distortion correction if reversed phase B0 images were available, and synB0-DisCo v3.0[48] if not.

### Tractography

Five different tractography methods were included: The standard clinical DTI-FACT based on manual white matter ROIs delineated[49] by 2 experienced operators on the Philips FiberTrack software was used as the reference method (FACT). Additionally, we included 4 automated approaches using probabilistic tensor-based tractography (TP)[50], and anatomically-filtered TP similar to ACT[32] (ATP), and probabilistic-CSD tractography using second order integration over fiber orientation distributions (iFOD2)[51], and anatomically-filtered iFOD2 (AiFOD2). This resulted in 5 different versions of every bundle of interest.

Automated tractography (TP, ATP, iFOD2, and AiFOD2) was done using the bundle-specific approach in KU Leuven Fun With Tracts v0.6 (KUL_FWT)[52] (https://github.com/KUL-Radneuron/KUL_FWT) with the addition of a model-based tractogram filtering step using streamline clustering (see supplementary methods). Automated tractography was done with 15,000(CST) and 6,000(AF) required number of streamlines for initial bundles. All generated tractograms were visually assessed for quality before further analysis. ANTs[42, 43] registration was used to bring all results to each subject’s T1 space for quantitative comparisons. Voxel-wise volumes for each bundle and bundle to TIV ratios were calculated per patient.

### DES coordinates processing and distance measures

Saved DES coordinates were exported from the neuronavigator and used to create spheres with 5mm radii[53] in FSL[41] in the same space as the anatomical image used during surgery, then warped to T1 space with ANTs[42, 43] for comparison with the tractography results. Minimum Euclidean distances were calculated between all DES coordinates and all voxels of the corresponding tractogram in Python v3.8. Distance measures were rounded up to integers (in mm) because increments smaller than a single voxel (1mm) were not considered meaningful.

### Statistical testing

#### Exploratory analysis

Distances between DES coordinates and tractograms can be considered akin to screening test results, and can thus be compared using confusion matrices, and ROC curves. However, due to the small sample size, and intersubject variation in number of DES coordinates, tractograms, and lesion size we opted for a more in-depth analysis. First **(RQ1)** we plotted the raw distance measures[54], then distances were averaged for each patient per DES response type to remedy the bias for patients with higher number of datapoints and ROC[55] curves were used to explore differences in sensitivity, specificity, and Youden’s index[56] ideal cutoffs.

Binary agreement/disagreement rates **(RQ2)** at different distance cutoffs were calculated and assessed using confusion matrices. Tractogram-DES pairs with distance less than the cutoff represented positive datapoints, true-positives if involving pDES, and false-positives if nDES, and those with distance above the cutoff represented negative datapoints, true-negatives if involving nDES and false-negatives if involving pDES.

#### Two-part linear modeling

Distance data thresholded at the cutoff determined by ROC of all pooled data were used for a two-part linear mixed model and posthoc testing to compare FT methods while correcting for differences in relative volumes of tractograms and lesions per patient. The thresholded distances data were a semicontinuous variable with excess zeros and an extremely right-skewed distribution, violating assumptions of normality. Therefore, and given the within-subjects nesting of repeated distance measures, we opted for a two-part model for longitudinal data[57, 58] **(RQ3)**. The model was estimated with the %MIXCORR macro provided by Tooze, Gunwald and Jones, 2002[58] and PROC NLMIXED in SAS studio v9.4 (SAS Institute, Cary, NC, USA).

The first part (A) predicted the probability of overlap (distance=0), and the second part (B) predicted the distances between nonoverlapping (distance>0) tractogram-DES coordinate pairs. Distances were the dependent variable and DES response, and tractography method (FACT, TP, ATP, iFOD2, and AiFOD2) were used as predictors in both parts of the model. Bundle-to-TIV and lesion-to-TIV ratios, were used as covariates of noninterest. Adaptive Hochberg’s[59] family-wise error-rate (FWE) correction was used to control for type(I) error in posthoc testing. Further detail can be found in supplementary material.

## Results

### Lesion segmentation, inpainting and tractography

Lesion masks had a median volume=44.70ml, minimum=1.20, maximum=232.129, and IQR=67.74ml. **Figure 2** shows the cumulative voxel-wise distribution of lesions in this sample of patients over the whole brain in MNI152 space. Lesion-inpainting and structural parcellation was successful in all patients. Tractography yielded 134 out of 135 attempted bundle reconstructions, with 1 failed right CST TP tractography (PT006). No bundles were excluded on inspection and none were repeated. **Figure 3 and S**.**figure 1** show all generated CST and AF tractograms, respectively. All methods showed a good level of visual agreement, but CSD-based methods resulted in more extensive tractograms. CSD captured the characteristic fanning of the CST into the lateral and inferior sensory-motor cortex, while DTI did not show this appearance, and AF CSD reconstructions reached further into the inferior frontal gyri and temporal lobes compared to DTI. As part of tractogram quality analysis we also explored the similarity of bundles generated with different FT methods, see supplementary material and **S.table 5** for details.

**Figure 2:**
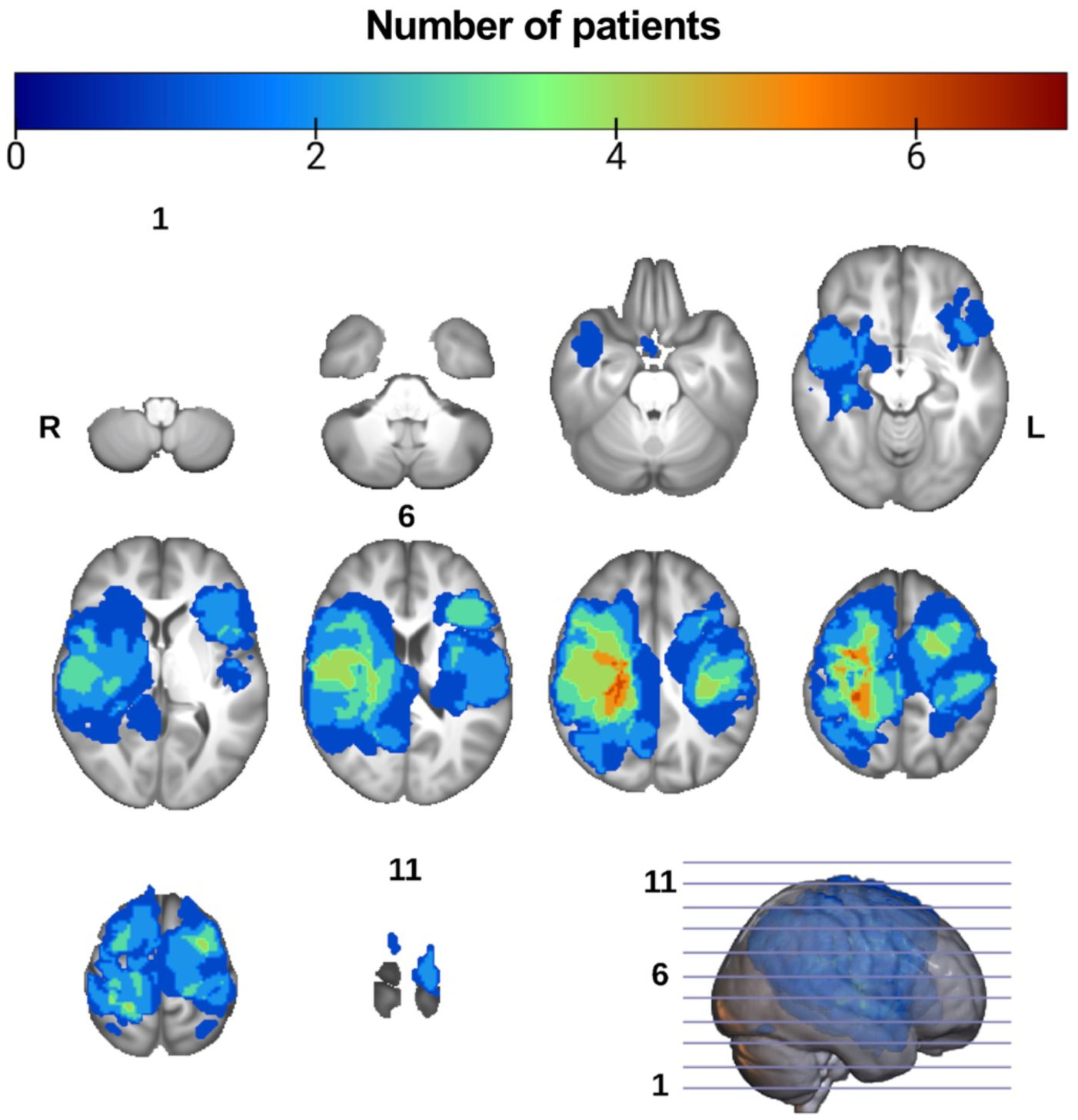
Spatial distribution of lesions from all patients overlaid onto the UK biobank T1 template brain in standard Montreal neurological institute (MNI) space. Perilesional edema was included in the masks of neoplasms if present. Overlay voxel intensities correspond to the sum of overlapping lesion masks from different patients. R = right, L = Left, slice numbers are indicated for the first, **middle and last slices.**

**Figure 3:**
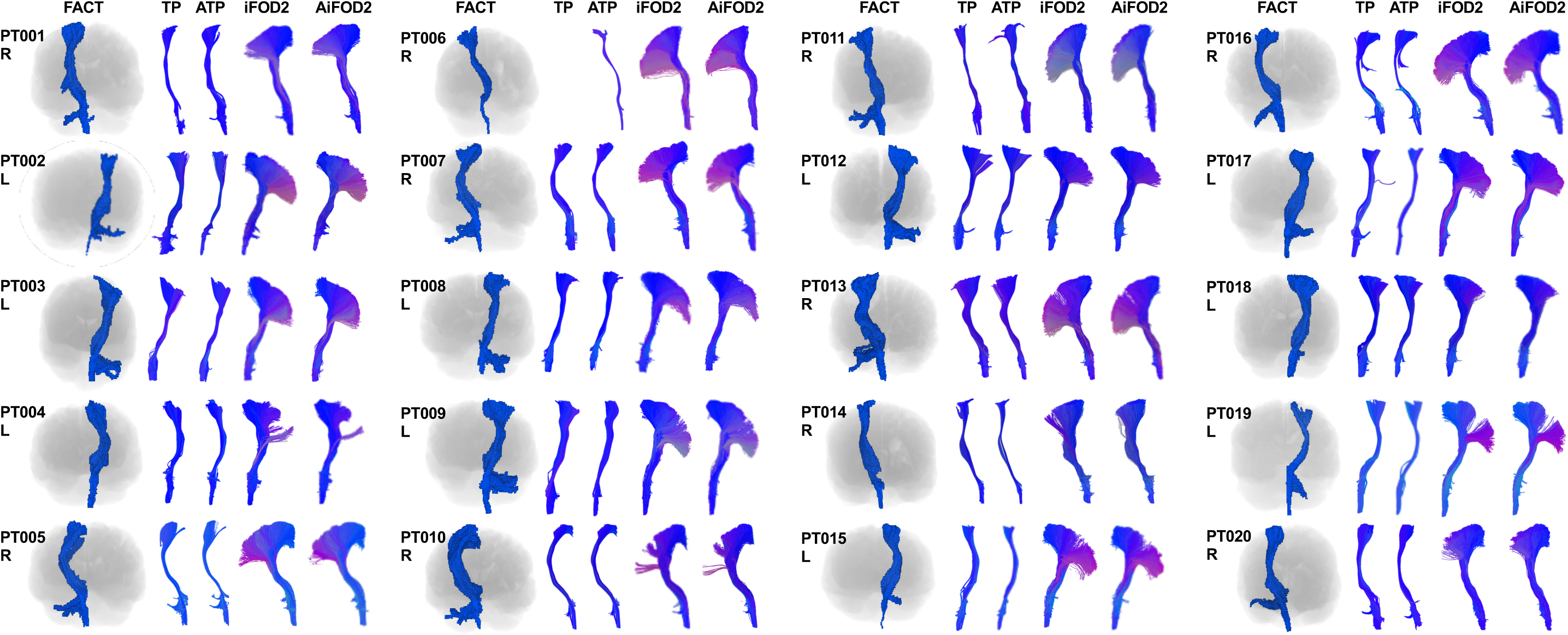
Corticospinal tractograms representative images from all methods in anterior view. The FACT tractogram outputs shown in blue are generated from volume rendered voxel masks with the T1-brain image silhouette shown underneath. The other 4 methods TP, ATP, iFOD2, and AiFOD2 are shown as 3D rendered streamlines with end-point directional color coding. All images are shown in radiological orientation. PT = patient, FACT = fiber assignment by continuous tracking, TP tensor probabilistic, ATP = anatomically constrained tensor probabilistic, iFOD2 = probabilistic tractography by second order integration over spherical harmonics, AiFOD2 = anatomically constrained iFOD2

### Intraoperative mapping and distance measures

DES mapping resulted in 51 pDES and 123 nDES coordinates, 3 patients had only nDES coordinates, namely PT011, PT012 and PT022, while 3 patients had only pDES coordinates, namely PT010, PT013, and PT015. **S.table 2** lists the DES tests, number of resulting pDES and nDES coordinates, elicited pDES response, acquired dMRI data, and bundle(s)-of-interest per patient. **Figure 4** shows images from 4 exemplar cases demonstrating the DES spheres and tractography results. Each tractogram was paired with pDES coordinates, and all nDES coordinates for distance measures, resulting in 860 distance measures with 174 measures for all methods except TP, which had 164 due to the failed right CST for PT006. **S.table 3** lists summarized descriptive statistics for distance measures, and **S.figure 2** shows the distribution of distances per bundle and DES response.

**Figure 4:**
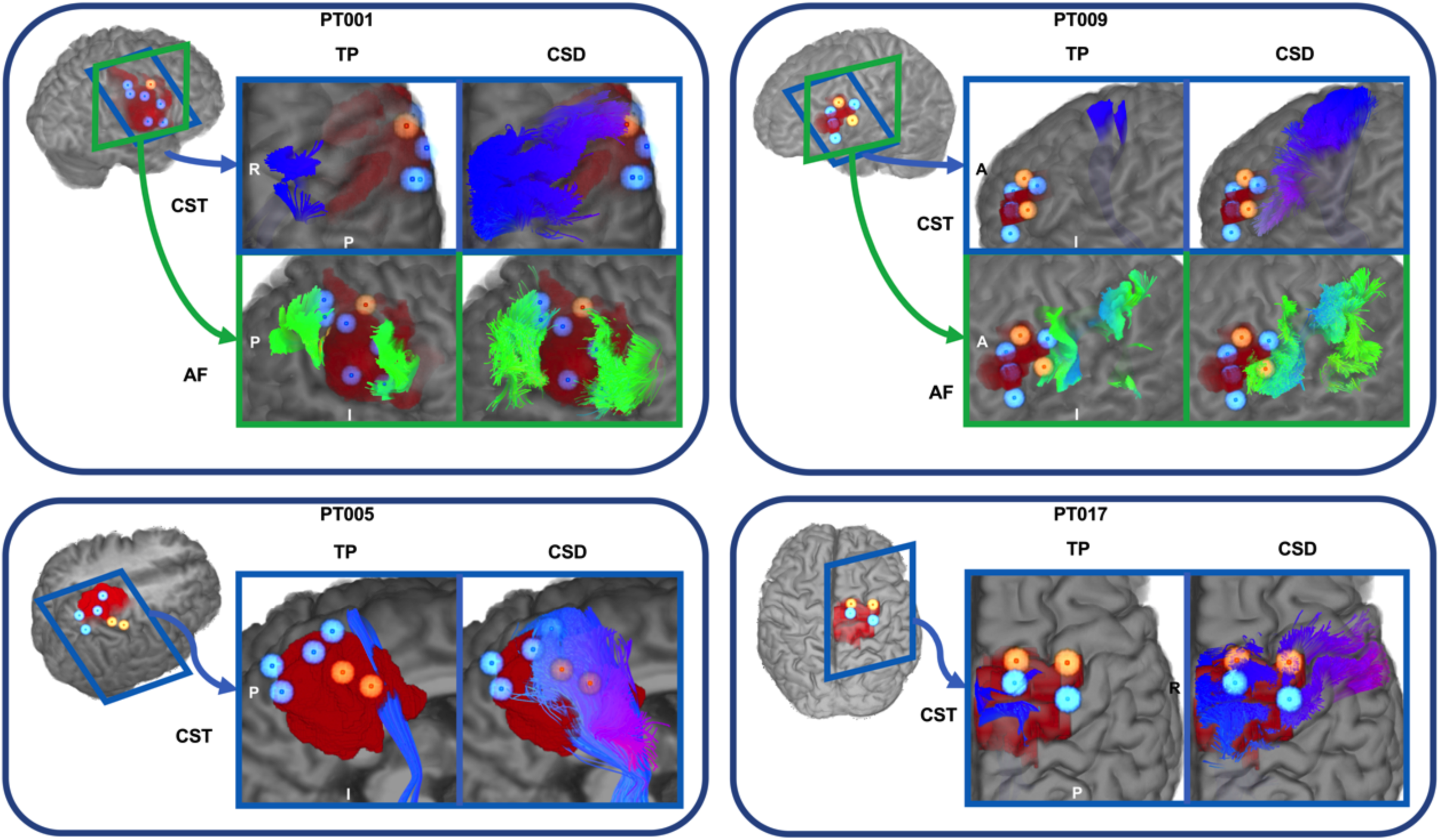
DES spheres, lesion masks and tractograms overlaid onto surface-rendered T1 images, for four example patients as generated with TP and iFOD2. Positive DES spheres are shown in semitransparent orange and their centers are shown in opaque orange, negative DES spheres are shown in semitransparent light blue with opaque blue centers. Tractograms are shown in endpoint-based directional color coding. TP = tensor probabilistic, iFOD2 = probabilistic tractography based on second order integration of spherical harmonic distributions, CST = corticospinal tract, AF = arcuate fasciculus, A = anterior, P = posterior, I = inferior, R = right

**Table 3:** Results of post hoc tests after two-part linear modeling for the CST at 12.5 mm distance cutoff.

|  |  |  |  |  |  |  |  |  |  |
| --- | --- | --- | --- | --- | --- | --- | --- | --- | --- |
| Test | Df | Model | T | P<sub>uncorr</sub> | P<sub>FWE</sub> | Model | T | P<sub>uncorr</sub> | P<sub>FWE</sub> |
| FACT v TP for DES and CST | 18 | A | -1.08 | 0.296 | 0.591 | B | -1.27 | 0.220 | 0.783 |
| FACT v ATP for DES and CST | 18 | A | -2.19 | 0.041 | 0.083 | B | -2.36 | 0.030 | 0.195 |
| FACT v iFOD2 DES and CST | 18 | A | 5.98 | <0.001 | <0.001 | B | 5.51 | <0.001 | <0.001 |
| FACT v AiFOD2 DES and CST | 18 | A | 5.62 | <0.001 | <0.001 | B | 6.42 | <0.001 | <0.001 |
| TP v ATP for DES and CST | 18 | A | -1.13 | 0.273 | 0.546 | B | -1.01 | 0.324 | 0.324 |
| TP v iFOD2 for DES and CST | 18 | A | 6.75 | <0.001 | <0.001 | B | 6.49 | <0.001 | <0.001 |
| TP v AiFOD2 for DES and CST | 18 | A | 6.46 | <0.001 | <0.001 | B | 7.43 | <0.001 | <0.001 |
| ATP v iFOD2 for DES and CST | 18 | A | 7.59 | <0.001 | <0.001 | B | 7.51 | <0.001 | <0.001 |
| ATP v AiFOD2 DES and CST | 18 | A | 7.32 | <0.001 | <0.001 | B | 8.49 | <0.001 | <0.001 |
| iFOD2 v AiFOD2 DES and CST | 18 | A | -0.44 | 0.665 | 0.665 | B | -0.27 | 0.451 | 0.451 |
| CST = corticospinal tract, FACT = fiber assignment by continuous tracking, TP = tensor probabilistic, ATP = anatomically constrained TP, iFOD2 = second order integration over fiber orientation distributions, AiFOD2 = anatomically constrained iFOD2, DF = degrees of freedom, T = t-statistic, P<sub>uncorr</sub> = uncorrected p values, P<sub>FWE</sub> = Hochberg family-wise error rate corrected p values |  |  |  |  |  |  |  |  |  |

### Statistical testing Exploratory analysis

The CST and AF tractograms tended to be closer to pDES than to nDES coordinates, regardless of FT method. CSD-based tractograms showed shorter distances than DTI-based tractograms to both nDES and pDES coordinates **(RQ2)**, see **S.figure 2** for plots of unthresholded distances. ROC curves for distance measures of all FT methods pooled together, as well as for both CSD-based and DTI-based FT methods are shown in **Figure 5**, and **S.figures 3** and **4** show the ROC curves for pooled raw distances, and each FT method separately with and without averaging.

**Figure 5:**
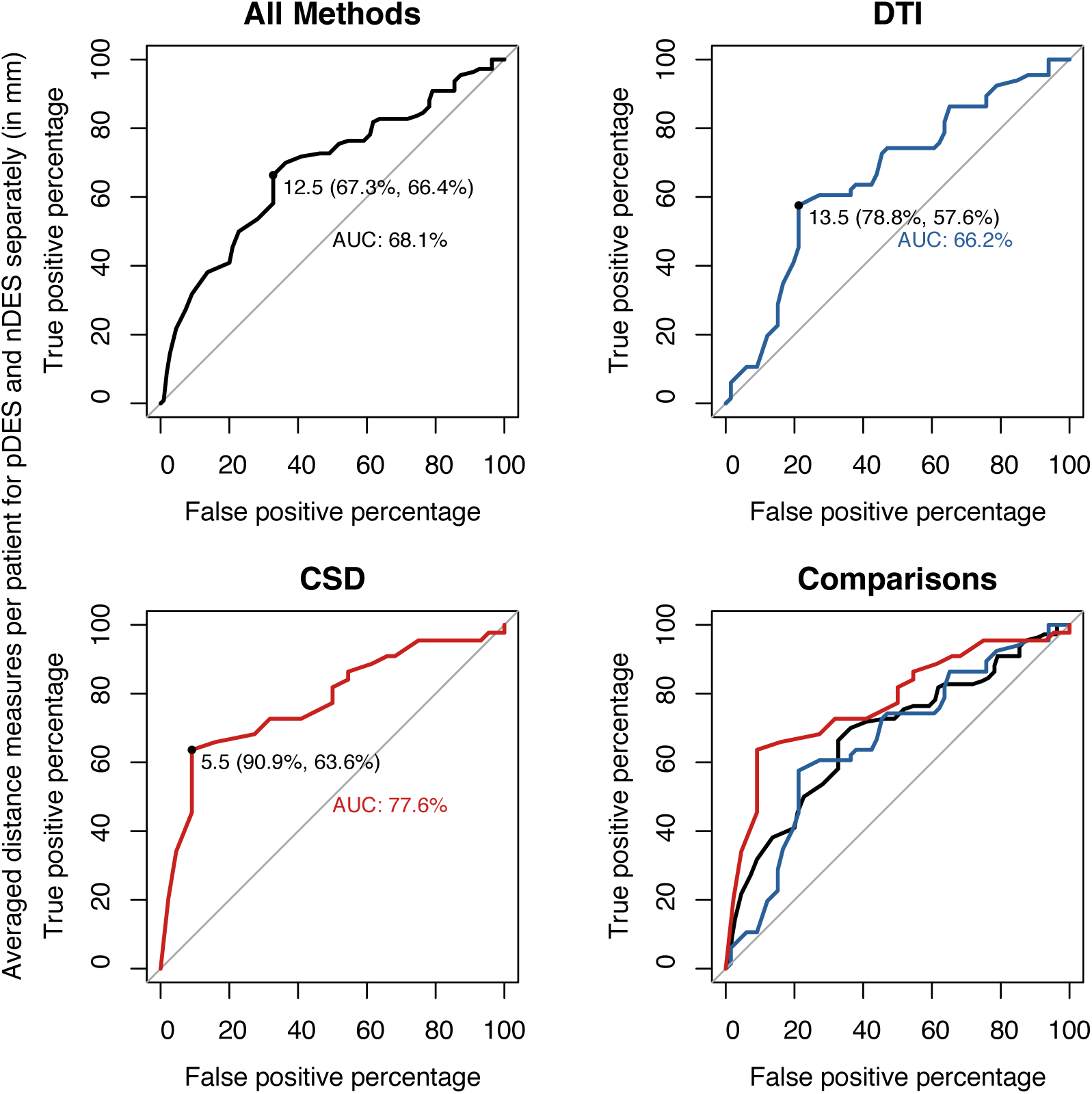
**ROC curves and optimal distance cutoffs** calculated using Youden’s method. Results for pooled data from all methods are shown in black, DTI (FACT, TP and ATP) in blue, and CSD (iFOD2 and AiFOD2) in red, FACT = fiber assignment by continuous tracking, TP tensor probabilistic, ATP = anatomically constrained tensor probabilistic, iFOD2 = probabilistic tractography by second order integration over spherical harmonics, AiFOD2 = anatomically constrained iFOD2

All ROC curves showed notable increase in sensitivity and comparatively smaller decrease in specificity for CSD-based compared to DTI-based FT, but there were no significant differences in AUC on pairwise DeLong tests, see **S.table 4**. Youden’s index determined distance cutoffs were also notably smaller for CSD-based methods (pooled=5.5mm, iFOD2=6.5mm, and AiFOD2=5.5mm) than DTI-based methods (pooled=13.5mm, FACT=19.5mm, TP=13mm and ATP=14.5mm), and the cutoff determined from all pooled data was 12.5mm. CSD-based methods were also associated with higher sensitivity at all ROC determined thresholds.

We used the 3 cutoffs determined from ROCs of pooled data of all methods, CSD, and DTI to define binary agreement/disagreement counts **(RQ2)** to compare FT methods using confusion matrices, see **Table 2**. These showed higher sensitivity and lower specificity for CSD-based compared to DTI-based tractography in both bundles at all distance cutoffs.

### Two-part model

The two-part linear model **(RQ3)** showed a significant main effect of tractography methods, and DES response type in both parts of the model. CSD-based methods (iFOD2 and AiFOD2) had larger probabilities of overlap (<12.5mm) and smaller distances (>12.5mm) compared to DTI-based methods (FACT, TP, ATP) for both pDES and nDES coordinates. Results also showed that pDES coordinates had significantly higher probability of overlap (T(18) = -7.70, p < 0.001), and were closer to tractograms than nDES coordinates were (T(18) = -5.09, p < 0.001). Bundle-to-TIV ratio had a significant effect (T(18) = -2.55, p = 0.020) only in the logistic part.

No significant interaction was found between DES response type and tractography method for the CST, indicating that differences between nDES and pDES were not significantly different between FT methods. Posthoc testing controlling for lesion-to-TIV and bundle-to-TIV ratios showed that CSD-based tractograms were more likely to overlap with and be closer to DES coordinates compared to DTI-based tractograms. These results are detailed in **Table 3** and **Figure 6, and S.figure 5** and **S.table 6** show results with 10.5mm distance cutoff.

**Figure 6:**
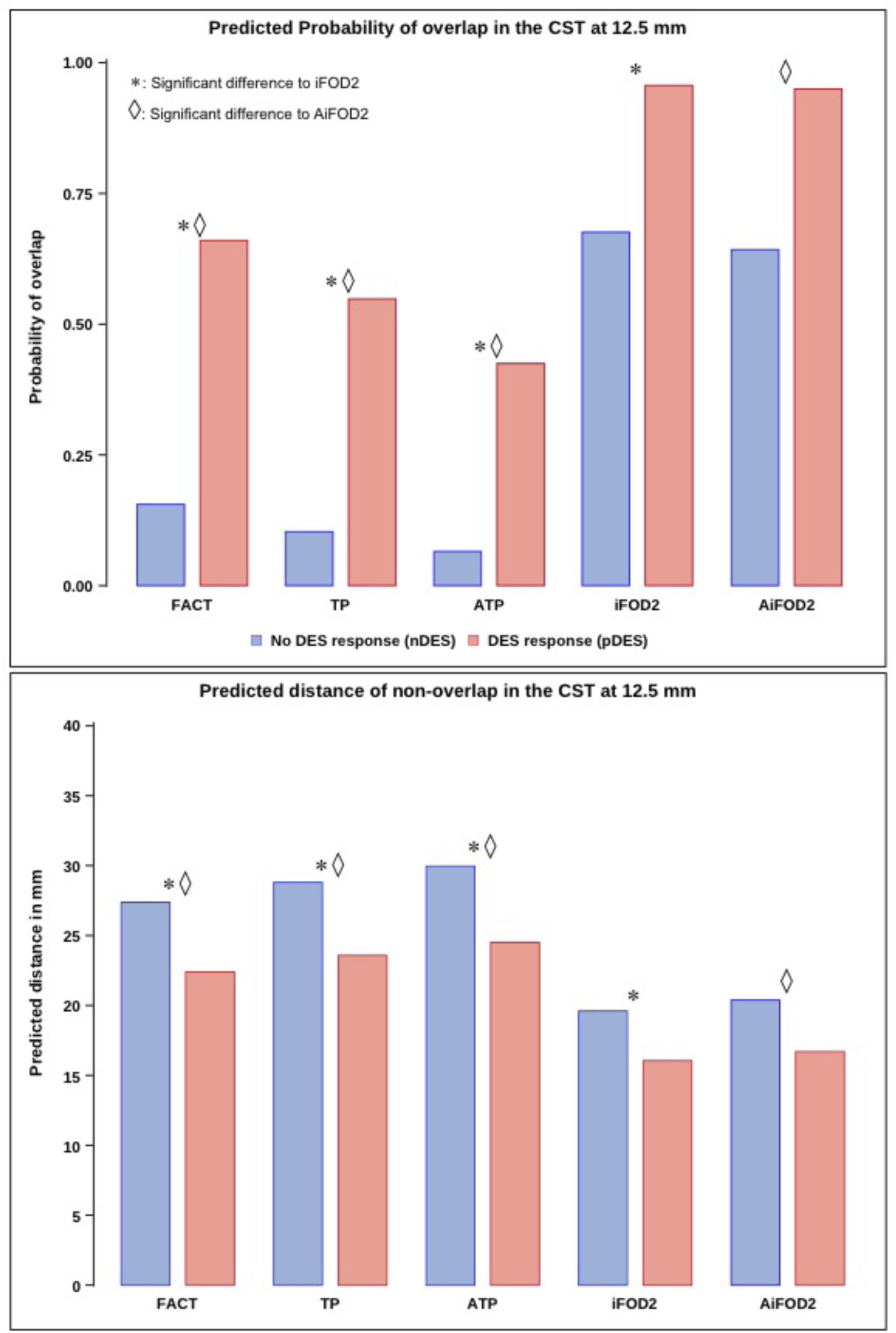
Bar plots for predicted probability of overlap between the CST and DES coordinates (Top) and predicted distances to nonoverlapping DES coordinates (Bottom) at 12.5 mm distance cutoff. CSD methods showed significantly higher probability of overlap, and lower distance if not overlapping compared to DTI methods. Differences between nDES and pDES were comparable between FT methods. CST = corticospinal tract, FACT = fiber assignment by continuous tracking, TP = tensor probabilistic, ATP = anatomically constrained TP, iFOD2 = probabilistic tractography by second order integration over spherical harmonics, AiFOD2 = anatomically constrained iFOD2

## Discussion

The primary aim of this work was to compare the accuracy of CSD and DTI tractography results as evaluated against intraoperative DES stimulation in a group of 22 preoperative neurosurgical patients. First, raw distance measures from CSD-based tractograms to DES coordinates were smaller than for DTI tractograms. These also showed that pDES coordinates were generally closer to tractograms compared to nDES coordinates. ROC curve plots and Youden’s index determined distance cutoffs, accuracy, sensitivity and specificity **(RQ1)** showed that pooled CSD methods (5.5mm) had a shorter distance cutoff compared to pooled DTI methods (13.5mm). This finding held with or without pooling and/or averaging. Overall higher sensitivity for CSD, and higher specificity for DTI tractograms was also found when comparing FT methods for their binary agreement/disagreement with DES at different distance cutoffs **(RQ2)** (5.5mm, 12.5mm, and 13.5mm).

The cutoff (12.5mm) determined by Youden’s index on the averaged all pooled data ROC was used to define binary agreement/disagreement (≤12.5/>12.5mm) rates between tractograms and DES. Lastly, we relied on a two-part linear mixed model and posthoc testing to investigate the effects of DES response, and differences between FT methods in probability of overlap(agreement) versus nonoverlap(disagreement), and distance measures for nonoverlapping tractogram-DES pairs while controlling for effects of variation in lesion and bundle volumes **(RQ3)**.

This showed that regardless of FT method, pDES coordinates were significantly closer and more probable to overlap with the CST compared to nDES coordinates. In other words, brain locations giving positive sensory-motor functional responses on stimulation (pDES), so called eloquent areas, were more likely to overlap with the CST, and if not overlapping they would typically closer than coordinates giving no functional effect on DES, so called non-eloquent. Furthermore, CST tractograms generated with CSD were significantly more probable to overlap with and be closer to DES coordinates compared to DTI, regardless of DES response. Posthoc testing confirmed that iFOD2 and AiFOD2 significantly outperformed FACT, TP and ATP in both parts of the model. These findings also indicate that the differences are not solely driven by larger volumes of CSD tractograms.

DTI-based tractography, when compared to DES, has previously been reported to have a generally satisfactory performance [11, 14, 60]. However, a closer look at the literature shows that these studies used manual tractography [61], and/or excluded negative DES coordinates from their analyses [62]. Additionally, the majority of previous work utilized comparatively simple statistical methods, and tended not to include covariates such as bundle volume, TIV or lesion volume in their analysis. In contrast to the literature, we a less than satisfactory performance for DTI-based FT. This, most likely, is because we included nDES data in the analysis, which exposes the problem with DTI FT, especially on smaller distance cutoffs.

Few studies thus far have compared different tractography methods to DES and/or DBS results so far [63–66], and all found a clear benefit from using more advanced tractography methods such as CSD. Our study adds to this growing body of evidence showing that advanced FT can improve the presurgical representation of white matter anatomy based on dMRI. This study also showed that manual and automated DTI FT performed comparably, indicating that automated methods relying on accurate structural parcellation adapted to effects of pathology can be comparable to relying on an expert user for manual FT, but that both performed worse than CSD FT as far distances and agreement with DES coordinates was concerned.

The mail limitation of this study is the small sample size, which motivated not including covariates for number of dMRI shells, pathology type, DES current, stimulator or approach used. However, the two-part linear model accounted for these factors by allowing for random intercepts and slopes per patient. Additionally, we did not include perioperative patient status in this analysis.

To summarize, both exploratory analyses and statistical modeling for the CST agreed that compared to DTI, CSD tractograms in this sample showed increased accuracy, with higher true-positives, at the cost of a smaller increase in false-positives, without significant reduction in true or false-negatives. No statistical testing was done for the AFs due to their small number (N=7) however it seemed to behave similar to the CST when evaluated qualitatively.

## Data Availability

All data produced in the present study are available upon reasonable request to the authors

