## Supplementary information for "A comparison of diffusion MRI presurgical tractography techniques with intraoperative mapping-based validation"

Radwan, A.M.[1,2], Emsell, L.[1,2,3,4], Vansteelandt, K.[2,3,4], Cleeren, E.[5], Peeters, R.[6], De Vleeschouwer, S.[2,7,8], Theys, T.[2,7,8], Dupont P.[2,9], Sunaert S.[1,2,6]

1. KU Leuven, Department of Imaging and pathology, Translational MRI, Leuven, Belgium
2. KU Leuven, Leuven Brain Institute (LBI), Department of Neurosciences, Leuven, Belgium
3. KU Leuven, Department of Neurosciences, Neuropsychiatry, Leuven, Belgium
4. KU Leuven, Department of Geriatric Psychiatry, University Psychiatric Center (UPC), Leuven, Belgium
5. UZ Leuven, Department of Neurology, Leuven, Belgium
6. UZ Leuven, Department of Radiology, Leuven, Belgium
7. KU Leuven, Department of Neurosciences, Research Group Experimental Neurosurgery and Neuroanatomy, Leuven, Belgium
8. UZ Leuven, Department of Neurosurgery, Leuven, Belgium
9. KU Leuven, Laboratory for Cognitive Neurology, Department of Neurosciences, Leuven, Belgium

### Tractogram filtering details

In addition to the methods previously described for KUL_FWT we also used template-based tractogram filtering with RecoBundles[1] as implemented in Scilpy [2] (scil_recognize_single_bundle.py) with model clustering threshold = 4 and pruning threshold = 8. Patient tractograms were warped to the template space prior to template-based filtering by applying ANTs generated warps and transforms to the bundle using MRtrix3.

### DES stimulation parameters

DES Stimulation parameters followed the protocol described by Duffau et al. [3] (60 Hz) in anaesthetized patients, and the low frequency protocol described by Zangaladze et al.[4] (5 Hz) in awake patients, stimulation started with 2-4 mA and gradually increased to 20 mA, or until a positive response was found. DES coordinates were considered positive if stimulation interfered with task performance or elicited a motor or sensory effect reported by the patient, observed by the attending neurophysiologist, or recorded on MEP/SSEP. DES coordinates were considered negative if no response could be elicited up to 20 mA in stimulation amplitude, and no effects were found if resection approaches its location.

Baseline cortical DES mapping was done before resection, testing locations were chosen based on visible anatomical landmarks in the surgical field and the coregistered MR images. DES tested locations were marked by sterile square markers, registered in the neuronavigation system, and continuously tested during resection. Coordinates were considered positive if an observable response was elicited, and negative if no response was elicited up to 20 mA nor during resection. Positive DES (pDES) and negative DES (nDES), cortical and subcortical coordinates, were saved and included in this analysis. The choice of DES mapping approach was based on lesion location and awake surgery feasibility. The bundles of interest were defined based on the neurosurgical treatment plan. **S.table 2** lists the details of the DES protocol per patient.

The DES spheres were collapsed to their centers of gravity (COG) after warping to T1 space of each patient and recreated with the same radii to mitigate deformations resulting from registration.

The following Python packages were used in an automated script in Python v3.8 to measure the minimum Euclidean distances between DES coordinates and tractograms: nibabel v3.2.2[5], numpy v1.22.3[6], scipy v1.4.1[7].

**Two-part linear model details**

The first part (A) used a logit-link for binary response (distance=0 vs. distance >0) and a generalized linear mixed model to predict probability of nonoverlap (distance > 0), and the second part (B) used a log-normal linear mixed model for the distance measures (distance >0) between nonoverlapping tractogram-DES coordinate pairs.

The thresholded distance measures were used as the dependent variable and DES response type (positive and negative), tractography methods (FACT, TP, ATP, iFOD2, and AiFOD2) were used as predictors in both parts of the model. Both models were adjusted for bundle to TIV and lesion to TIV ratios, which were used as covariates, and DES response type x tractography methods interactions were considered but were omitted as they were not significant.

For ease of interpretation, we discuss and plot the probability of overlap (distance = 0) for the first part (A), and log distances (B) from the second part are back-transformed to distance in mm.

Results were interpreted in the following context: Predicted probability of overlap (distance < cutoff) with pDES coordinates was analogous to true-positive rate. Predicted probability of overlap with nDES coordinates was analogous to false-positive rate. Predicted distances to non-overlapping (distance > cutoff) nDES coordinates were analogous to true-negative rate, and predicted distances to non-overlapping pDES coordinates were analogous to false-negative rate.

| Supplementary table 1(a): Patient demographics, pathology results, lesion lobe, side and volume | | | | | | | | |
|---|---|---|---|---|---|---|---|---|
| **Patients** | **Age** | **Gender** | **Lesion type** | **WHO grade** | **Pathology report** | **Lesion lobe** | **Lesion side** | **Lesion volume (ml)** |
| **PT001** | 60 - 65 | M | Glioma | IV | Glioblastoma | Fronto-parietal | R | 66.809 |
| **PT002** | 5 – 10 | F | FCD | I | Type I | Frontal | L | 1.200 |
| **PT003** | 35 – 40 | M | Meningio ma | | Transitional type meningioma | Frontal | L | 54.079 |
| **PT004** | 30 – 35 | F | Glioma | II | Oligodendrogliom a | Fronto-parietal | L | 124.775 |
| **PT005** | 70 – 75 | M | Glioma | IV | Glioblastoma | Parieto-occipital | R | 32.382 |
| **PT006** | 40 – 45 | F | Glioma | IV | Glioblastoma | Temporo-fronto-parietal | R | 217.077 |
| **PT007** | 65 – 70 | F | Glioma | IV | Glioblastoma | Frontal | R | 26.779 |
| **PT008** | 35 – 40 | M | Glioma | IV | Glioblastoma | Temporo-fronto-parietal | L | 88.917 |
| **PT009** | 10 – 15 | M | FCD | | Type IIB | Frontal | L | 15.666 |
| **PT010** | 60 – 65 | M | Glioma | IV | Glioblastoma | Fronto-parietal | R | 93.831 |
| **PT011** | 40 – 45 | M | Glioma | II | Multifocal astrocytoma | Fronto-temporo-parieto-occipital | R | 123.293 |
| **PT012** | 60 – 65 | F | Glioma | II | Oligodendrogliom a | Frontal | L | 53.331 |
| **PT013** | 45 – 50 | F | FCD | | Type IIB | Parietal | L | 5.224 |
| **PT014** | 30 – 35 | M | Glioma | III | Oligodendrogliom a | Frontal | R | 92.154 |
| **PT015** | 35 – 40 | M | Glioma | IV | Glioblastoma | Frontal | R | 232.129 |
| **PT016** | 30 – 35 | F | Glioma | II | Oligodendrogliom a | Parietal | L | 22.553 |
| **PT017** | 60 – 65 | M | Glioma | IV | Glioblastoma | Parietal | R | 42.517 |
| **PT018** | 30 – 35 | F | Glioma | III | Oligodendrogliom a | Frontal | L | 46.887 |
| **PT019** | 55 – 60 | M | Glioma | III | Astrocytoma | Fronto-temporal | L | 40.208 |
| **PT020** | 55 – 60 | M | Glioma | II | Oligodendrogliom a | Frontal | L | 11.531 |
| **PT021** | 15 – 20 | M | FCD | | Type IIB | Frontal | R | 4.806 |
| **PT022** | 10 – 15 | M | DNET | I | DNET | Frontal | L | 34.207 |
| PT = patient, M = male, F = female, FCD = focal cortical dysplasia, DNET = dysembryoplastic neuroepithelial tumor, WHO = world health organization, R = right, L = left, ml = milliliters, IQR = interquartile range | | | | | | | | |

| Supplementary table 1(b): Summarized demographics, pathological type, distribution and volumes | | | | |
|---|---|---|---|---|
| **Age and Gender** | **Lesion** | | | |
| | **Type** | **Side** | **Cerebral lobar distribution** | **Lesion volumes** |
| Age range=8 - 73 years<br>Median age=39.5 years<br>IQR=28 years<br>14 males<br>8 females | 18 neoplasms:<br>16 gliomas<br>1 meningioma<br>1 dysembryoplastic neuro-epithelial tumor<br>4 focal cortical dysplasia | 10 R<br>12 L | 3 fronto-parietal<br>1 fronto-temporal<br>11 frontal<br>3 parietal<br>1 parieto-occipital<br>2 Temporo-fronto-parietal<br>1 Fronto-temporo-parieto-occipital (multifocal lesion) | Range=1.2 - 232.13 ml<br>Median=44.70<br>IQR=71.74 |
| PT = patient, M = male, F = female, FCD = focal cortical dysplasia, DNET = dysembryoplastic neuroepithelial tumor, WHO = world health organization, R = right, L = left, ml = milliliters, IQR = interquartile range | | | | |

| Supplementary table 2: Direct electrical stimulation (DES) mapping and dMRI and tractography details | | | | | | |
|---|---|---|---|---|---|---|
| **Patients** | **Awake surgery** | **DES +ve/-ve** | **Positive DES effect** | **Stimulation threshold Ctx/SubCtx** | **dMRI data** | **Bundles** |
| **PT001** | Y | 1/6 | Motor dysarthria | 20 / -mA | 2 shells + RP-B0 | AF + CST |
| **PT002** | N | 4/6 | Motor right leg, foot, wrist, and hand | 6 / 4 mA | 2 shells + RP-B0 | CST |
| **PT003** | Y | 3/3 | Motor right leg, foot, and hand | 4 mA | 2 shells + RP-B0 | CST |
| **PT004** | Y | 1/7 | Motor face, right hand, and arm | - / 5 mA | 1 shell (b2500) + RP-B0 | AF + CST |
| **PT005** | Y | 2/4 | Motor left hand | 16 / 5 mA | 2 shells + RP-B0 | CST |
| **PT006** | Y | 2/8 | Sensory-Motor left leg | 20 / 5 mA | 2 shells + RP-B0 | CST |
| **PT007** | Y | 2/6 | Sensory-motor left leg | 16 / 10 mA | 2 shells + RP-B0 | CST |
| **PT008** | Y | 2/5 | Difficulty finding words, and sensory mouth | 20 / 2 mA | 1 shell (b1200) | AF + CST |
| **PT009** | Y | 2/4 | Speech arrest, paraphasia, and motor face | 8 / - mA | 2 shells + RP-B0 | AF + CST |
| **PT010** | N | 3/0 | Motor left leg | Not recorded | 2 shells + RP-B0 | CST |
| **PT011** | Y | 0/6 | None | - | 2 shells + RP-B0 | CST |
| **PT012** | Y | 0/7 | None | - | 1 shell (b1200) + RP-B0 | AF |
| **PT013** | Y | 7/0 | Sensory-motor right arm and hand | 10 / 5 mA | 2 shells + RP-B0 | CST |
| **PT014** | Y | 2/6 | Motor left hand and foot | 12 / - mA | 2 shells + RP-B0 | CST |
| **PT015** | N | 2/0 | Motor left hand | - / 10 mA | 2 shells + RP-B0 | CST |
| **PT016** | Y | 3/6 | Motor right hand | 12 / 5 mA | 1 shell (b1200) + RP-B0 | CST |
| **PT017** | Y | 2/2 | Motor left foot, leg, hand and arm | 4 / 12 mA | 2 shells + RP-B0 | CST |
| **PT018** | Y | 3/8 | Motor right hand, lips, and dysarthria | 20 / 5 mA | 1 shell (b1200) | CST |
| **PT019** | Y | 1/6 | Motor right hand | 20 / 10 mA | 2 shells + RP-B0 | CST |
| **PT020** | Y | 1/4 | Motor right arm | - / 10 mA | 1 shell (b1200) + RP-B0 | AF + CST |
| **PT021** | Y | 3/6 | Motor left wrist and hand | 4 mA | 2 shells + RP-B0 | CST |
| **PT022** | Y | 0/8 | None | - | 2 shells + RP-B0 | AF |

PT = patient, DES = direct electrical stimulation, +ve = positive DES, -ve = negative DES, dMRI = diffusion magnetic resonance imaging, AF = arcuate fasciculus, CST = corticospinal tract, Y = awake surgery, N = general anaesthesia with motor and somatosensory evoked potentials (MEP/SSEP), Ctx = cortical, SubCtx= subcortical, mA = milliampere, RP-B0 = reversed-phase non-diffusion weighted spin-echo EPI volume.

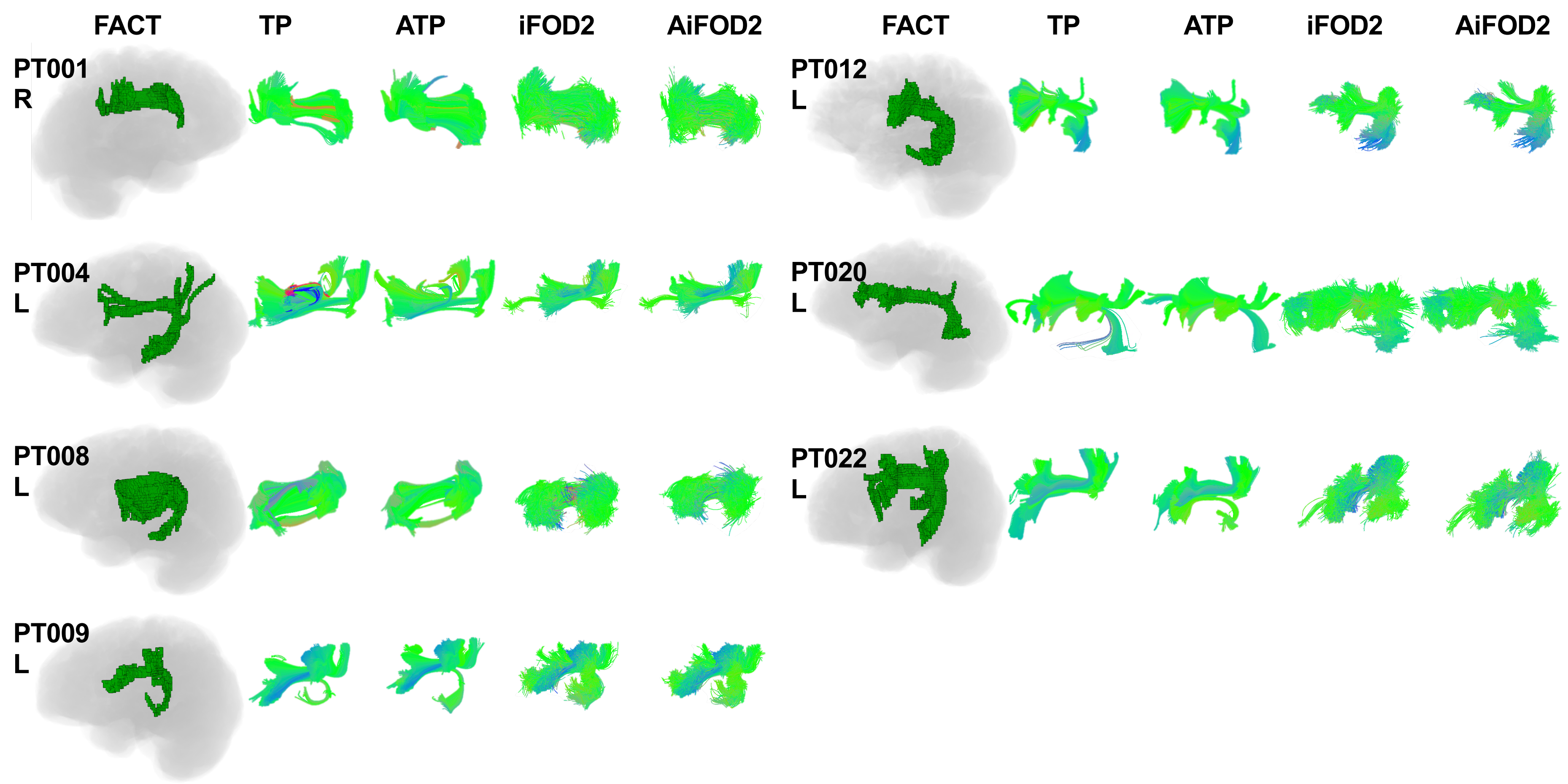

**Supplementary figure 1 Arcuate fasciculi (AF) representative images** using all methods in lateral projection. The FACT tractogram outputs shown in green are generated from volume rendered voxel masks with the T1-brain image silhouette shown underneath. The other 4 methods TP, ATP, iFOD2, and AiFOD2 are shown as 3D rendered streamlines with end-point directional color coding. PT = patient, FACT = fiber assignment by continuous tracking, TP tensor probabilistic, ATP = anatomically constrained tensor probabilistic, iFOD2 = probabilistic tractography by second order integration over spherical harmonics, AiFOD2 = anatomically constrained iFOD2

| Supplementary table 3: Descriptive statistics for distance measures per tractography method | | | | | | | | | | | |
|---|---|---|---|---|---|---|---|---|---|---|---|
| **DES type** | **Bundle** | **FT** | **Mean** | **StDev** | **Median** | **Q1** | **Q3** | **IQR** | **Var** | **Range** | **N** |
| **pDES** | **CST** | **FACT** | 12.37 | 11.73 | 8 | 2.25 | 21 | 18.75 | 137.57 | 0 - 39 | 46 |
| | | **TP** | 13.14 | 11.76 | 8.50 | 3.75 | 22 | 18.25 | 138.21 | 0 - 39 | 44 |
| | | **ATP** | 14.78 | 11.45 | 10.50 | 5.25 | 23 | 17.75 | 131.15 | 0 - 39 | 46 |
| | | **iFOD2** | 5.22 | 6.75 | 2 | 1 | 6 | 5 | 45.51 | 0 - 34 | 46 |
| | | **AiFOD2** | 4.89 | 6.06 | 3 | 1 | 6 | 5 | 36.68 | 0 - 27 | 46 |
| **nDES** | | **FACT** | 27.24 | 12.47 | 28 | 19 | 37 | 18 | 155.42 | 3 - 57 | 93 |
| | | **TP** | 27.37 | 12.18 | 29 | 19.50 | 37 | 17.50 | 148.40 | 1 - 54 | 85 |
| | | **ATP** | 30.23 | 12.16 | 32 | 22.50 | 39 | 16.50 | 147.83 | 3 - 58 | 93 |
| | | **iFOD2** | 15.68 | 10.79 | 13 | 6.50 | 22 | 15.50 | 116.42 | 0 - 43 | 93 |
| | | **AiFOD2** | 15.78 | 10.71 | 13 | 8 | 21 | 13 | 114.59 | 0 - 48 | 93 |
| **pDES** | **AF** | **FACT** | 12.80 | 6.18 | 13 | 12 | 17 | 5 | 38.20 | 3 - 19 | 5 |
| | | **TP** | 9.40 | 3.85 | 10 | 6 | 12 | 6 | 14.80 | 5 - 14 | 5 |
| | | **ATP** | 8.60 | 3.58 | 8 | 6 | 10 | 4 | 12.80 | 5 - 14 | 5 |
| | | **iFOD2** | 5.60 | 5.41 | 3 | 2 | 8 | 6 | 29.30 | 1 - 14 | 5 |
| | | **AiFOD2** | 5.20 | 4.82 | 4 | 2 | 8 | 6 | 23.20 | 0 - 12 | 5 |
| **nDES** | | **FACT** | 15 | 10.71 | 12 | 6.25 | 21.75 | 15.50 | 114.76 | 0 - 40 | 30 |
| | | **TP** | 13.90 | 11.26 | 13.50 | 3.50 | 21 | 17.50 | 126.78 | 0 - 42 | 30 |
| | | **ATP** | 14 | 11.84 | 11.50 | 4.25 | 20.50 | 16.25 | 140.07 | 0 - 42 | 30 |
| | | **iFOD2** | 9.37 | 7.79 | 8.50 | 2.75 | 12.75 | 10 | 60.66 | 0 - 33 | 30 |
| | | **AiFOD2** | 9.67 | 8.25 | 8 | 3.50 | 13.50 | 10 | 68.09 | 0 - 35 | 30 |

DES = direct electrical stimulation, pDES = positive DES, nDES = negative DES, FT = fiber tractography method, StDev = standard deviation, Q = quartile, IQR = interquartile range, Var = variance, N = number, FACT = fiber assignment by continuous tracking, TP = tensor probabilistic, ATP = anatomically constrained TP, iFOD2 = probabilistic tractography by second-order integration over spherical harmonics, AiFOD2 = anatomically constrained iFOD2

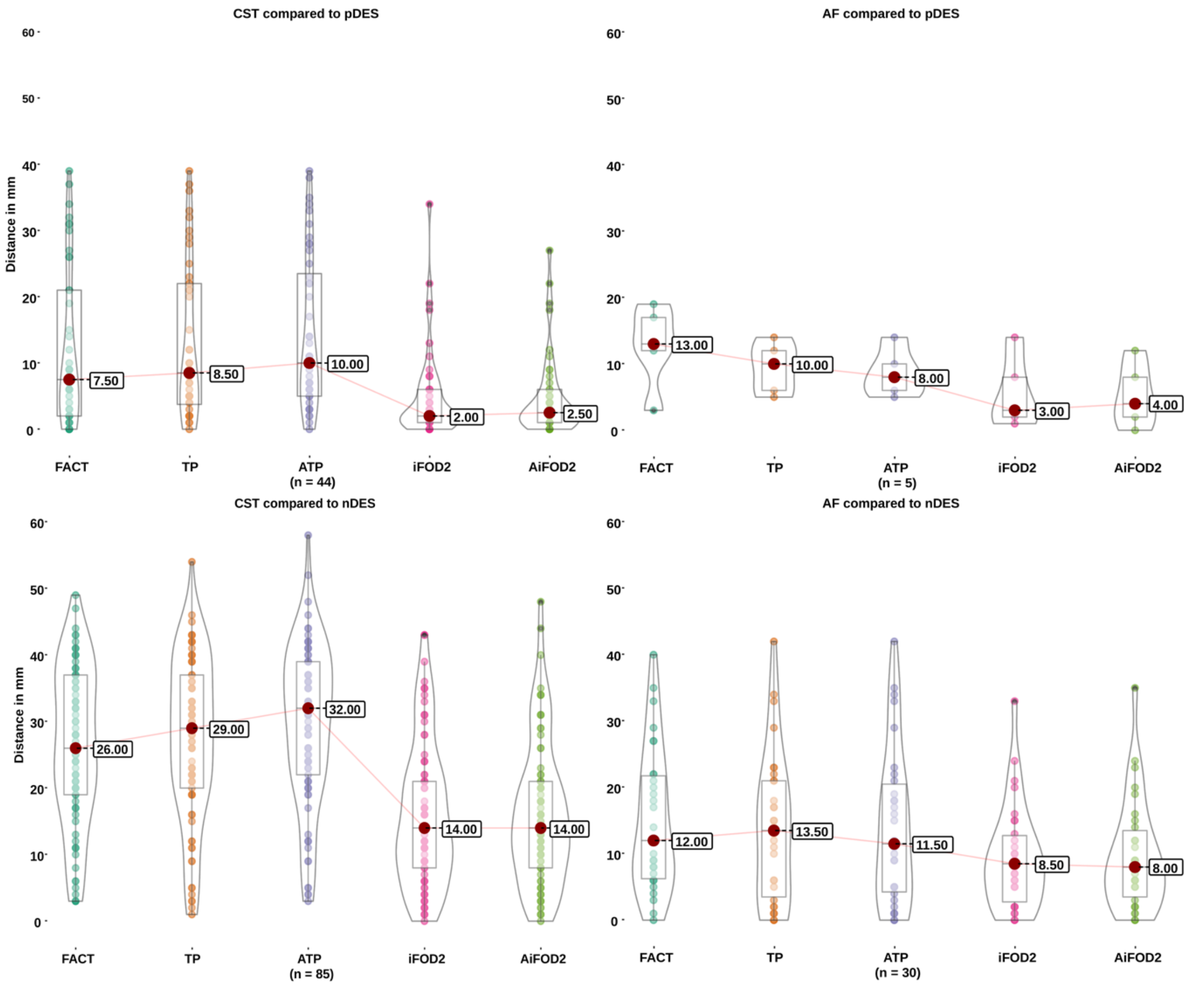

**Supplementary figure 2 Box and violin plots of distance measures by bundle type, DES response type and tractography method.** Results for the CST are shown on the left, for the AF on the right, for pDES on top, and for nDES on the bottom. The highlighted circle at the center of each violin plot indicates the median distance value, the box within each violin indicates the interquartile range, and the central line represents the range. FACT = fiber assignment by continuous tracking, TP tensor probabilistic, ATP = anatomically constrained tensor probabilistic, iFOD2 = probabilistic tractography by second order integration over spherical harmonics, AiFOD2 = anatomically constrained iFOD2

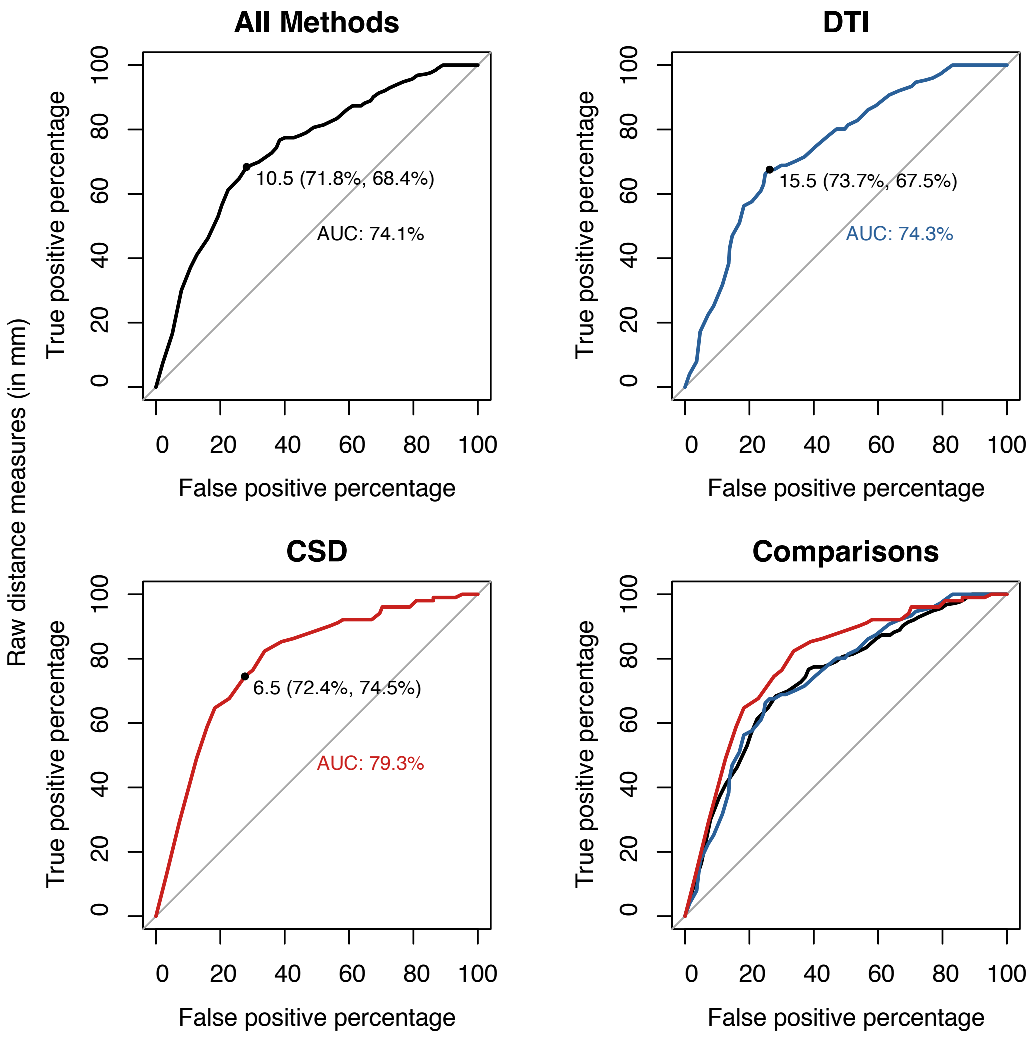

**Supplementary figure 3** ROC plots for raw distance measures pooled for all methods (black), DTI (blue), and CSD (red), showing the optimal distance cutoff, and area under the curve (AUC), and comparisons in bottom right panel. FACT = fiber assignment by continuous tracking, TP tensor probabilistic, ATP = anatomically constrained tensor probabilistic, iFOD2 = probabilistic tractography by second order integration over spherical harmonics, AiFOD2 = anatomically constrained iFOD2

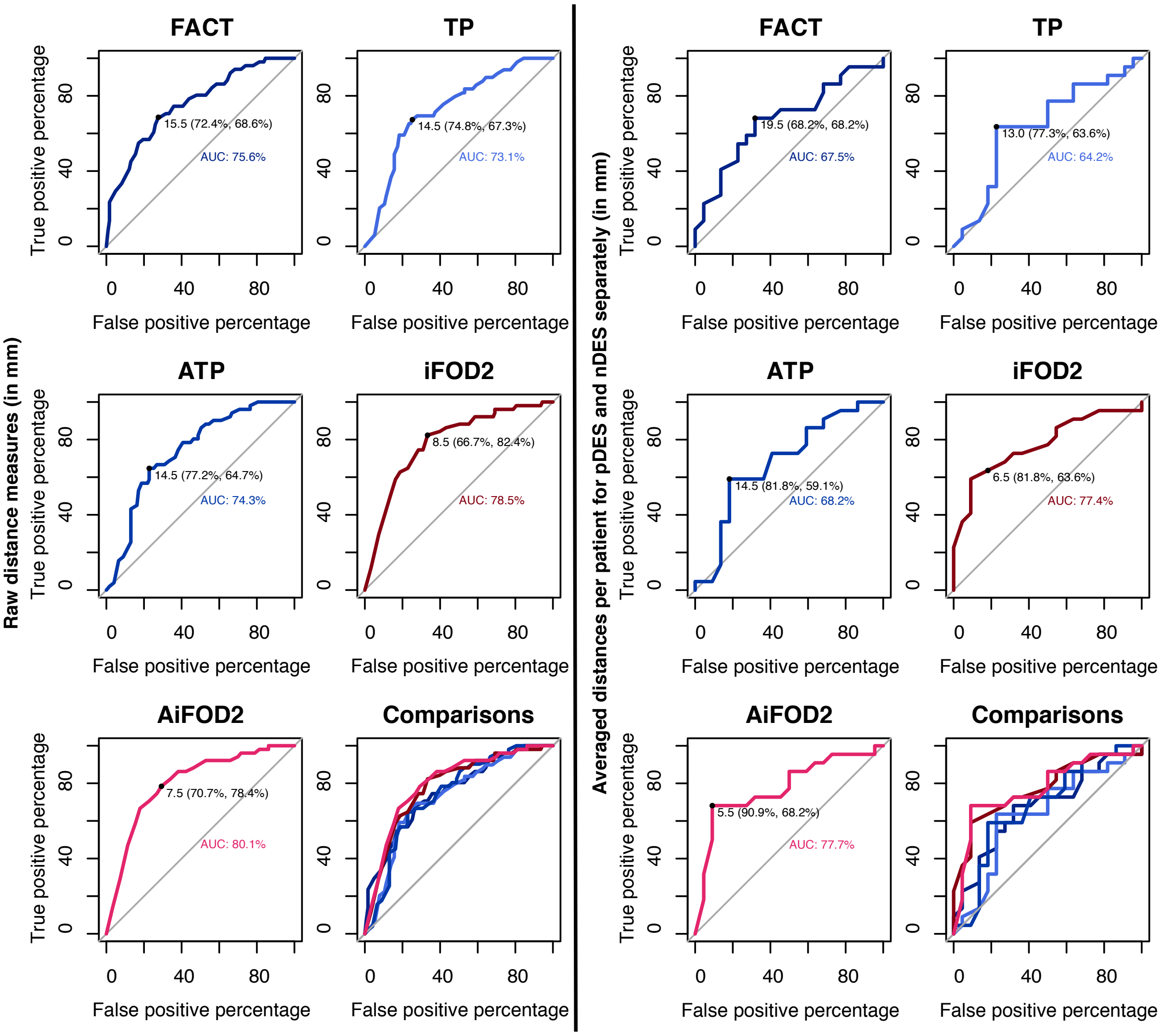

**Supplementary figure 4**: ROC plots for each tractography method using raw distances on the left, and using averaged distances per patient for nDES and pDES separately on the right. Also shown are the optimal distance cutoffs, and area under the curve (AUC), and all methods ROCs compared in bottom right panel. FACT = fiber assignment by continuous tracking, TP tensor probabilistic, ATP = anatomically constrained tensor probabilistic, iFOD2 = probabilistic tractography by second order integration over spherical harmonics, AiFOD2 = anatomically constrained iFOD2

| Supplementary table 4: Summarized results of DeLong pairwise tests comparing the ROC curves from averaged distance measures | | | | | | |
|---|---|---|---|---|---|---|
| **Pairwise comparisons** | **Estimate1** | **Estimate2** | **Statistic** | **p.value** | **Conf.low** | **Conf.high** |
| **FACT v TP** | 67.46 | 64.15 | 0.62 | 0.54 | -0.07 | 0.14 |
| **FACT v ATP** | 67.46 | 68.18 | -0.22 | 0.82 | -0.07 | 0.06 |
| **FACT v iFOD2** | 67.46 | 77.38 | -1.45 | 0.15 | -0.23 | 0.03 |
| **FACT v AiFOD2** | 67.46 | 77.69 | -1.55 | 0.12 | -0.23 | 0.03 |
| **TP v ATP** | 64.15 | 68.18 | -0.81 | 0.42 | -0.14 | 0.06 |
| **TP v iFOD2** | 64.15 | 77.38 | -1.84 | 0.07 | -0.27 | 0.01 |
| **TP v AiFOD2** | 64.15 | 77.69 | -1.92 | 0.05 | -0.27 | 0.00 |
| **ATP v iFOD2** | 68.18 | 77.38 | -1.37 | 0.17 | -0.22 | 0.04 |
| **ATP v AiFOD2** | 68.18 | 77.69 | -1.46 | 0.15 | -0.22 | 0.03 |
| **iFOD2 v AiFOD2** | 77.38 | 77.69 | -0.21 | 0.84 | -0.03 | 0.03 |
| ROC = receiver operating characteristic, Conf. = confidence interval, DES = direct electrical stimulation, pDES = positive DES, nDES = negative DES, FT = fiber tractography method, StDev = standard deviation, Q = quartile, IQR = interquartile range, Var = variance, N = number, FACT = fiber assignment by continuous tracking, TP = tensor probabilistic, ATP = anatomically constrained TP, iFOD2 = probabilistic tractography by second-order integration over spherical harmonics, AiFOD2 = anatomically constrained iFOD2 | | | | | | |

#### Tractogram similarity analysis results (46 words)

DSC and JI scores were used to evaluate the similarity of tractograms across different FT methods. As expected, we found higher similarity between AiFOD2 and iFOD2 than between TP, ATP and iFOD2 bundles. However, clinical DTI FACT tractograms also showed higher DSC and JI compared to TP and ATP, see **S.table 5** for summarized descriptive statistics of tractogram similarity measures. Tractogram shape similarity analysis using DSC and JI with iFOD2 tractograms as the reference showed the expected pattern of mild to moderate similarity between DTI and iFOD2 tractograms. This also showed that the CSD-based and DTI-based tractograms were not identical among themselves, confirming the differences noted visually.

| Supplementary table 5: Summarized descriptive statistics for bundle similarity measures for all methods compared to iFOD2 | | | | | | | | |
|---|---|---|---|---|---|---|---|---|
| **Tractography methods** | **FACT** | | **TP** | | **ATP** | | **AiFOD2** | |
| **Measures** | **JI** | **DSC** | **JI** | **DSC** | **JI** | **DSC** | **JI** | **DSC** |
| **max** | 0.41 | 0.59 | 0.34 | 0.51 | 0.42 | 0.59 | 0.76 | 0.86 |
| **min** | 0.10 | 0.18 | 0.13 | 0.23 | 0.06 | 0.11 | 0.51 | 0.68 |
| **mean** | 0.26 | 0.41 | 0.24 | 0.39 | 0.20 | 0.32 | 0.66 | 0.80 |
| **median** | 0.25 | 0.39 | 0.24 | 0.39 | 0.20 | 0.33 | 0.69 | 0.82 |
| **stdev** | 0.09 | 0.11 | 0.06 | 0.08 | 0.08 | 0.11 | 0.07 | 0.05 |
| **IQR** | 0.08 | 0.10 | 0.06 | 0.08 | 0.04 | 0.06 | 0.02 | 0.02 |

iFOD2 = second order integration over fiber orientation distributions, FACT = fiber assignment by continuous tracking, TP = tensor probabilistic, ATP = anatomically constrained TP, AiFOD2= anatomically constrained iFOD2, JI = Jaccard index, DSC = Dice similarity coefficient, stdev = standard deviation, IQR = interquartile range

| Supplementary table 6: Results of post hoc tests after two-part linear modeling for the CST at 10.5 mm distance cutoff | | | | | | | | | |
|---|---|---|---|---|---|---|---|---|---|
| **Test** | **Df** | **Model** | **T** | **$P_{uncorr}$** | **$P_{FWE}$** | **Model** | **T** | **$P_{uncorr}$** | **$P_{FWE}$** |
| **FACT v TP for DES and CST** | 18 | A | -1.07 | 0.298 | 0.595 | B | -1.28 | 0.218 | 0.537 |
| **FACT v ATP for DES and CST** | 18 | A | -2.30 | 0.034 | 0.102 | B | -2.48 | 0.023 | 0.070 |
| **FACT v iFOD2 DES and CST** | 18 | A | 5.01 | <0.001 | <0.001 | B | 7.19 | <0.001 | <0.001 |
| **FACT v AiFOD2 DES and CST** | 18 | A | 4.98 | <0.001 | <0.001 | B | 7.26 | <0.001 | <0.001 |
| **TP v ATP for DES and CST** | 18 | A | -1.26 | 0.225 | 0.595 | B | -1.14 | 0.269 | 0.537 |
| **TP v iFOD2 for DES and CST** | 18 | A | 5.94 | <0.001 | <0.001 | B | 8.24 | <0.001 | <0.001 |
| **TP v AiFOD2 for DES and CST** | 18 | A | 5.93 | <0.001 | <0.001 | B | 8.33 | <0.001 | <0.001 |
| **ATP v iFOD2 for DES and CST** | 18 | A | 6.85 | <0.001 | <0.001 | B | 9.44 | <0.001 | <0.001 |
| **ATP v AiFOD2 DES and CST** | 18 | A | 6.84 | <0.001 | <0.001 | B | 9.54 | <0.001 | <0.001 |
| **IFOD2 v AiFOD2 DES and CST** | 18 | A | -0.06 | 0.956 | 0. 956 | B | -0.07 | 0.941 | 0.941 |

CST = corticospinal tract, FACT = fiber assignment by continuous tracking, TP = tensor probabilistic, ATP = anatomically constrained TP, iFOD2 = second order integration over fiber orientation distributions, AiFOD2 = anatomically constrained iFOD2, DF = degrees of freedom, T = t-statistic, $P_{uncorr}$ = uncorrected p values, $P_{FWE}$ = Hochberg family-wise error rate corrected p values

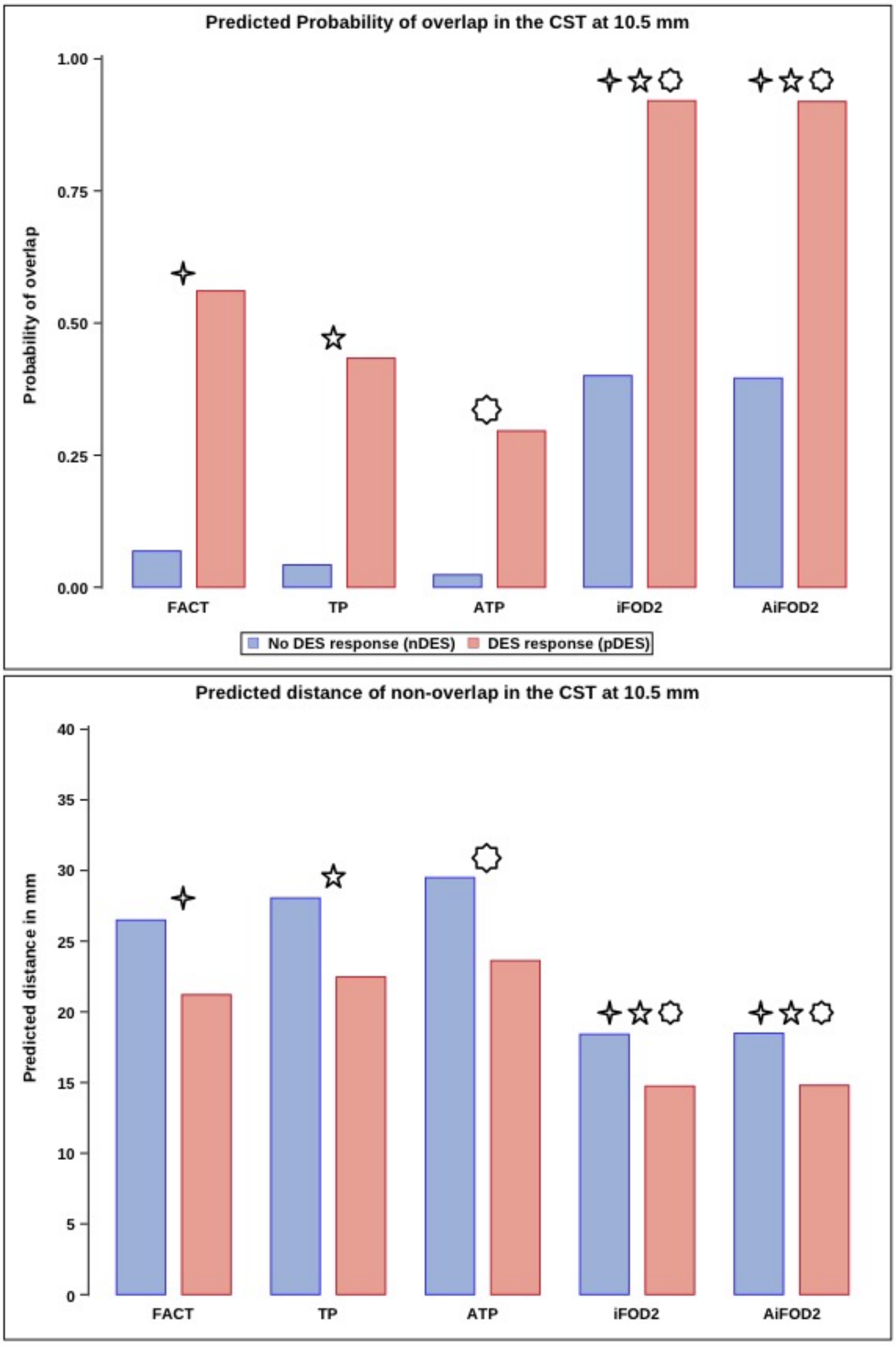

**Supplementary figure 5:** Bar plots for predicted probability of overlap between the CST and DES coordinates (Top) and predicted distances to nonoverlapping DES coordinates (Bottom) at 10.5 mm distance cutoff. CSD methods showed significantly higher probability of overlap, and lower distance if not overlapping compared to DTI methods. Differences between nDES and pDES were comparable between FT methods. CST = corticospinal tract, FACT = fiber assignment by continuous tracking, TP = tensor probabilistic, ATP = anatomically constrained TP, iFOD2 = probabilistic tractography by second order integration over spherical harmonics, AiFOD2 = anatomically constrained iFOD2, four-, five-, and eight-pointed stars denote significant pairwise differences between FT methods.
